# Association of polygenic risk scores for coronary artery disease with subsequent events amongst established cases

**DOI:** 10.1101/19009431

**Authors:** Laurence J. Howe, Frank Dudbridge, A. Floriaan Schmidt, Chris Finan, Spiros Denaxas, Folkert W. Asselbergs, Aroon D. Hingorani, Riyaz S. Patel

## Abstract

**Background:** There is growing evidence that polygenic risk scores (PRS) can be used to identify individuals at high lifetime risk of coronary artery disease (CAD). Whether they can also be used to stratify risk of subsequent events among those surviving a first CAD event remains uncertain.

**Methods:** Using two subsamples of UK Biobank, defined at baseline as prevalent CAD (N=10,287) and without CAD (N=393,108), we evaluated associations between a CAD PRS and incident cardiovascular and fatal outcomes, during a median follow up of 7.8 years.

**Results:** A 1 S.D. higher PRS was associated with increased risk of incident MI in participants without CAD (OR 1.33; 95% C.I. 1.29, 1.38), but the effect estimate was markedly attenuated in those with prevalent CAD (OR 1.15; 95% C.I. 1.06, 1.25); heterogeneity P =0.0012. Additionally, among prevalent CAD cases, we found evidence of an inverse association between the CAD PRS and risk of all-cause death (OR 0.91; 95% C.I. 0.85, 0.98) compared to those without CAD (OR 1.01; 95% C.I. 0.99, 1.03); heterogeneity P =0.0041. A similar inverse association was found for ischaemic stroke (Prevalent CAD (OR 0.78; 95% C.I. 0.67, 0.90); without CAD (OR 1.09; 95% C.I. 1.04, 1.15), heterogeneity P <0.001).

**Conclusions:** Bias induced by case stratification and survival into UK Biobank may attenuate, or reverse, associations of polygenic risk scores derived from case-control studies or populations initially free of disease. Polygenic risk scores for subsequent events should be derived from new genome wide association studies conducted in patients with established disease.

**Key messages:**

- CAD PRS are positively associated with incident myocardial infarction risk amongst established CAD cases.
- However, the effect size is attenuated compared to estimates from CAD-free populations.
- CAD PRS are inversely associated with mortality and stroke risk amongst established CAD cases.
- These associations may reflect index event bias induced by stratifying on case status.
- Dedicated GWAS of coronary disease progression are required to improve prediction of subsequent event risk.

## Introduction

Coronary artery disease (CAD) is heritable, with over 300 independent genetic loci with additive effects known to influence disease risk having been identified in Genome-Wide Association Studies (GWAS) (1–4). Exploiting the increasing amount of risk variation captured by identified loci, recent studies have illustrated the potential use of CAD polygenic risk scores (PRS) for identifying individuals at elevated risk of CAD (5–7), where the scores are based on counts of the number of risk alleles carried.

However, the extent to which CAD PRS derived from general population cohorts or case-control GWAS are associated with CAD disease progression, as characterized by subsequent or recurrent events amongst diseased cases, remains unclear. Indeed, established risk variants for onset of CAD may not necessarily equate with variants influencing risk of subsequent events because of genuine aetiological differences between the pathophysiology of the two states (8) (9). Alternatively, even if variants influencing disease onset also genuinely influence progression, associations may be distorted because of index event bias, where conditioning on an index event (e.g. presence of CAD) may induce confounded associations between risk factors in the sample of individuals with the index event (10, 11). The lack of a strong association between the major CAD risk locus at 9p21 and subsequent event risk highlights possible differences in genetic associations of CAD risk variants dependent on case status (12) (13).

Previous studies have found some evidence that CAD PRS are associated with increased risk of subsequent events (e.g. recurrent MI and revascularization) (7, 14–19), although a recent study in a French-Canadian population found that CAD PRS are substantially less effective at predicting recurrence and incident cases than prevalence (7). Stronger conclusions have been limited by the modest sample sizes of recurrence studies, with most previous studies including less than 5000 cases, as well as the inconsistency of cardiovascular and fatal endpoints across different studies.

Using two subsamples of UK Biobank, defined as individuals (a) free of CAD; and (b) those with evidence of prevalent CAD at enrolment, we aimed to evaluate the extent to which associations between CAD genetic risk variants and incident events differ when restricting to a case-only sample while also running several exploratory analyses to detect and account for potential index event bias.

## Methods

### Data sources

#### UK Biobank

UK Biobank is a large-scale cohort study, which recruited approximately 500,000 individuals aged between 40 and 69 years from across the United Kingdom. Genotype data are available for the majority of participants with extensive phenotype data collected via questionnaire at baseline. Study participants are linked to electronic health record data from Hospital Episode Statistics (HES), secondary care data containing International Classification of Diseases, 10^th^ Revision (ICD10) and Office of Population Censuses and Surveys Classification of Surgical Operations (OPCS) codes relating to study participants diagnoses and operative procedures. Study participants are also linked to the mortality register from the Office of National Statistics (ONS) which contains data on death, time of death, as well as primary and secondary causes (20).

For the purposes of this study, we used a sample of 408,480 individuals which was generated by starting with the full sample and removing individuals of non-European descent, individuals with more than 10 putative third-degree relatives in the kinship table and individuals who were flagged in quality control (sex mismatch, heterozygosity and individual missingness). We then defined two subsamples for our analyses; a) baseline CAD controls, generated by removing prevalent CAD cases (see below) and individuals that self-reported as having had a heart attack (Field ID: 6150-0.0, 20002-0.0) or coronary angioplasty/coronary artery bypass grafts (Field ID: 20004-0.0), and b) prevalent CAD cases identified using the following ICD10 (I21-I25, Z955) and OPCS codes (K40-K46, K471, K49, K50, K75) from HES occurring before their study enrolment date. In secondary analyses, CAD cases were stratified into coronary artery disease with myocardial infarction (CADMI) cases (ICD10: I21-23, I241, I252) and coronary artery disease cases with no evidence of myocardial infarction (CADnoMI).

Phenotype data collected at baseline included sex, age, body mass index (BMI) (Field ID: 21001-0.0), systolic blood pressure (SBP) (Field ID: 4080-0.0), self-reported type II diabetes (Field ID: 2443-0.0), self-reported smoking status (Field ID: 20116-0.0) and self-reported statin use (Field ID: 20003).

Incident events after recruitment into the UK Biobank were ascertained using ICD10 and OPCS codes from HES using similar codes to published phenotyping algorithms (21). Incident cardiovascular events of interest included: MI (I21-23, I241, I252), heart failure (I110, I130, I132, I260, I50), ischemic stroke (I63, I693), stroke (I60-64, I69) and revascularization (K40-46, K471, K49, K50, K75). Fatal events of interest included cardiovascular (CVD) death, CAD death and all-cause death and were ascertained using primary cause of death from mortality register data using ICD codes for cause-specific mortality. Composite events of interest were combined CAD death/MI and a combined variable including all cardiovascular outcomes. More information on relevant ICD10 and OPCS codes are contained in **Supplementary Table 1**.

UK Biobank study participants (N=488,347) were assayed using the UK BiLEVE Axiom™ Array by Affymetrix1 (N= 49,950) and the closely related UK Biobank Axiom™ Array (N= 438,427). Directly genotyped variants were pre-phased using SHAPEIT3 (22) and imputed using Impute4 and the UK10K (23), Haplotype Reference Consortium (24) and 1000 Genomes Phase 3 (25) reference panels with post-imputation data including ∼96 million genetic variants (26, 27).

#### CARDIOGRAMPlusC4D

CARDIOGRAMPlusC4D (28) is a global collaboration of studies using a case-control approach to identify genetic variants associated with the presence of CAD. In this study, we used publicly available GWAS summary data from a recent consortium study independent of UK Biobank (2), which were downloaded from the CARDIOGRAM website (http://www.cardiogramplusc4d.org/data-downloads/).

### Statistical analysis

#### CAD PRS

We used GWAS summary data from CARDIOGRAMPlusC4D to construct a CAD PRS of SNPs. Initially, all SNPs meeting a P-value inclusion criterion (P<5×10^−6^) were considered in order to generate a restrictive score containing only loci with relatively strong evidence for association with CAD. Highly correlated markers were then removed by LD clumping (R^2^<0.2, 250 kb distance threshold) the summary data using PLINK v1.9 (29) and the 1000 Genomes Phase 3 (GBR samples) (25). The final CAD PRS included 182 SNPs with the contribution of each SNP weighted by the GWAS effect estimates.

#### CAD PRS and incident events

Associations between the CAD PRS and incident cardiovascular (stroke, ischemic stroke, myocardial infarction, heart failure, revascularization), fatal (all-cause death, CVD death and CAD death) and composite (all CVD, CAD death or MI) outcomes were evaluated in the baseline CAD case and control samples. Logistic regression was used to estimate associations, with all analyses adjusted for age and sex. For comparison, we presented effect estimates in the two samples and tested for heterogeneity between these estimates (30). As a sensitivity analysis, we stratified the case only sample by type of CAD (CADMI / CADnoMI) and compared estimates between the two samples with a test for heterogeneity. All estimates were presented in terms of a standard deviation increase in the PRS.

#### CAD PRS and baseline covariates

Index event bias may lead to confounded associations between different CAD risk factors (e.g. between CAD PRS and BMI) amongst cases which are not present or are not as strong in samples of CAD free individuals. In turn, these may confound associations between risk factors and subsequent events (31). Therefore, we quantified and compared associations between CAD risk factors and the CAD PRS in the case and CAD-free samples.

As covariates of interest we chose established risk factors for CAD available in UK Biobank (age, sex, SBP, BMI, type II diabetes, ever smoking and statin use), which were collected at study baseline. Linear or logistic regression models in R v3.6.0 were used to estimate associations between the CAD PRS and covariates in the baseline CAD case and control samples. Analyses with age and sex as the phenotypes of interest were run unadjusted, with all other regression models including age and sex as covariates. For comparison, we presented the value of covariates of interest at quintiles (20%, 40%, 60%, 80%) of the CAD PRS distribution and formally tested for heterogeneity between estimates for first and subsequent events (30).

#### Accounting for index event bias

To evaluate the potential effects of index event bias on our analyses, we ran sensitivity analyses using two different approaches. First, we repeated the CAD PRS and incident events analyses in the baseline CAD case sample, including SBP, BMI, type II diabetes, ever smoking and statin use as covariates. These CAD risk factors were included as covariates to attempt to account for confounded associations between the CAD PRS and these covariates relating to index event bias.

Second, we used a recently proposed method to correct for index event bias in GWAS. SNP effects on prognosis (i.e. on events occurring after an index event) are adjusted using residuals from the regression of the SNP effects on the index event against the SNP effects on prognosis. The main caveat with the approach is that it assumes that the direct genetic effects on incidence and prognosis are independent (32).

In this instance, the index event is existing CAD so we used GWAS summary data from the CARDIOGRAMPlusC4D GWAS (2). For a GWAS of prognosis, we used the UK Biobank CAD case sample (N=10,287) to perform a GWAS of all-cause mortality using a logistic model in snptest v2.5.2 (33), including age, sex and the first ten principal components as covariates. As suggested previously (32), we then extracted 116,438 independent SNPs common to both the CARDIOGRAMPlusC4D and the UK Biobank GWAS summary statistics by restricting to well-imputed SNPs (INFO>0.99) and LD pruning (250 kb step window, 5 SNP step size, r2=0.1) using the 1000 Genomes GBR samples (Phase 3) (25) as a reference panel. These SNPs were then used to calculate the slope correction estimate using the SIMEX (34) option in the IndexEvent.R package with a Hedges-Olkin estimate calculated as a sensitivity analysis. When applying this in practice, the SIMEX slope estimates did not converge using all SNPs, even with 10,000 simulations, so we decided to reduce noise by removing SNPs not strongly associated with the index trait and calculate the slope using a subset of 5564 SNPs which were nominally associated with the index trait in CARDIOGRAMPlusC4D (P<0.05).

Next, we adjusted the betas and standard errors in the UK Biobank GWAS of mortality using the slope of the regression. For example, the adjusted betas were calculated by subtracting the product of the slope estimate and the CARDIOGRAM incidence beta estimate from the prognosis beta for each SNP. Finally, to estimate the association between the CAD PRS and mortality amongst CAD cases from summary data (instead of individual level data as previously), we used an inverse-variance weighted method (35) (36) across 54 independent SNPs (28) using CAD as the exposure and mortality as the outcome. The subset of chosen independent SNPs reached genome-wide significance in the largest GWAS independent of UK Biobank. Estimates were presented in terms of the association of an increase in the CAD PRS, corresponding to an odds increase of CAD, with log-odds of mortality. For comparison, we estimated the PRS association before and after correction.

## Results

### CAD PRS and incident events

Associations between the PRS and incident events differed greatly between the two samples, with confidence intervals non-overlapping for eight out of the ten outcomes tested (heterogeneity P < 0.05). In the CAD free sample, we found strong evidence of positive associations between the CAD PRS and incident cardiovascular and fatal outcomes such as MI (OR 1.34; 95% C.I. 1.29, 1.38), CAD death (OR 1.31; 95% C.I 1.23, 1.40) and ischemic stroke (OR 1.09; 95% C.I. 1.04, 1.15). In contrast, in the prevalent CAD sample, we found evidence of a positive, but attenuated association with MI (OR 1.15; 95% C.I. 1.06, 1.25; Int P=0.012), weak evidence for an association with CAD death (OR 0.96; 95% C.I. 0.85, 1.08; Int P=9.1×10^−6^) and evidence of inverse associations with all-cause death (OR 0.91; 95% C.I. 0.85, 0.98; Int P = 0.0041) and ischemic stroke (OR 0.78; 95% C.I. 0.67, 0.90; Int P = 1.8×10^−5^) (**Figure 1** / **Table 1**). Amongst prevalent CAD cases, we did not find strong evidence of heterogeneity by CAD subtype (CAD without prior MI, CAD with prior MI) with overlapping confidence intervals across all outcomes (**Supplementary Table 2**).

**Table 1:** Associations between CAD GRS and incident events.

| **Outcome** | **No CAD at baseline**<br>**(N=393,108)**<br>**OR (95% C.I.)** | **Prevalent CAD cases**<br>**(N=10,287)**<br>**OR (95% C.I.)** | **Heterogeneity P-value** |
| --- | --- | --- | --- |
| CAD death/MI | 1.33 (1.29, 1.38) | 1.11 (1.03, 1.20) | $1.4 \times 10^{-5}$ |
| CV death | 1.20 (1.14, 1.26) | 0.97 (0.87, 1.09) | 0.0012 |
| All-cause death | 1.01 (0.99, 1.03) | 0.91 (0.85, 0.98) | 0.0041 |
| CAD death | 1.31 (1.23, 1.40) | 0.96 (0.85, 1.08) | $9.1 \times 10^{-6}$ |
| MI | 1.34 (1.29, 1.38) | 1.15 (1.06, 1.25) | 0.0012 |
| Revasc | 0.99 (0.97, 1.01) | 0.91 (0.84, 1.00) | 0.067 |
| Heart failure | 1.00 (0.93, 1.08) | 0.97 (0.86, 1.10) | 0.67 |
| Stroke | 1.05 (1.01, 1.09) | 0.88 (0.78, 1.00) | 0.0094 |
| Ischaemic stroke | 1.09 (1.04, 1.15) | 0.78 (0.67, 0.90) | $1.8 \times 10^{-5}$ |
| All CVD | 1.06 (1.04, 1.08) | 0.99 (0.93, 1.04) | 0.013 |

<sup>†</sup> All OR per 1 S.D. increase in CAD GRS of 182 SNPs

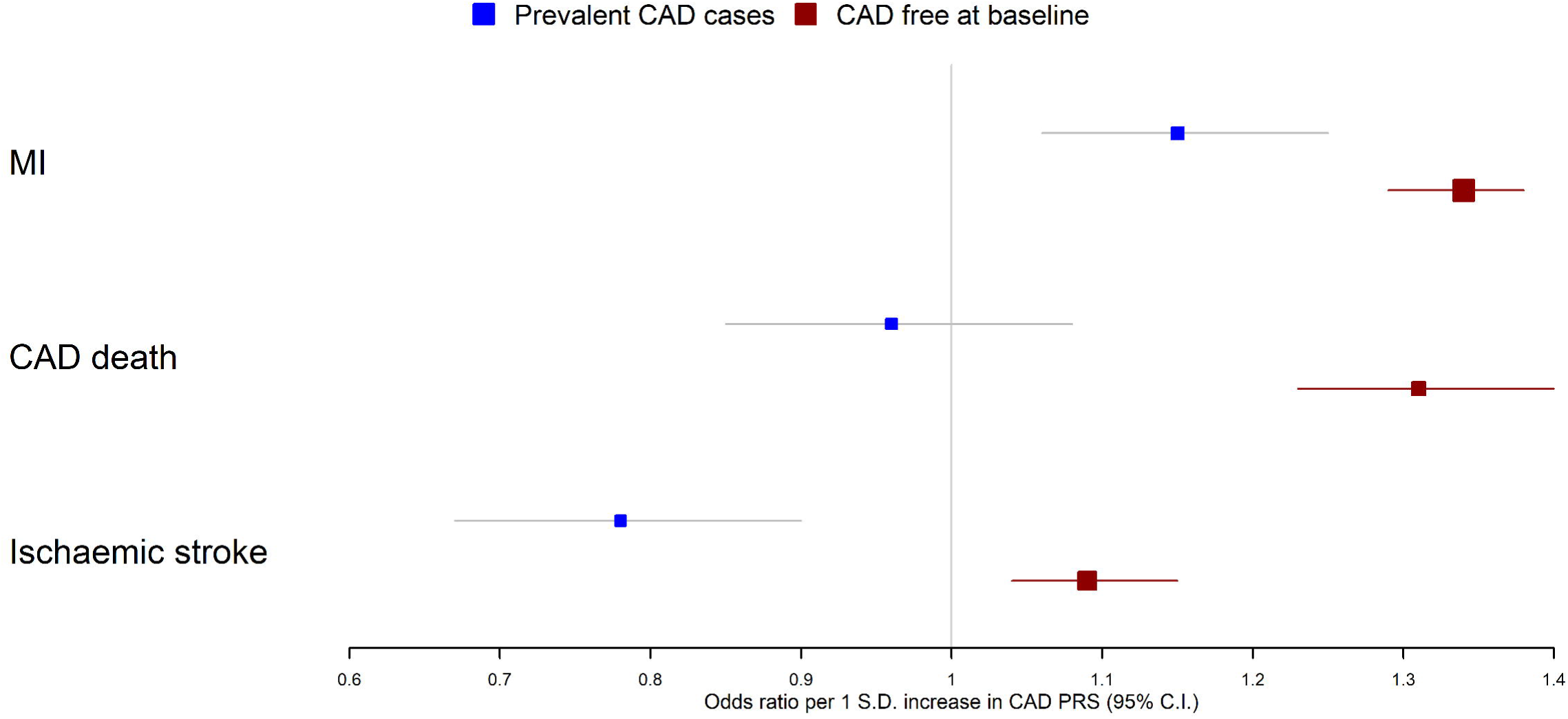

### CAD PRS and baseline covariates

The CAD PRS was inversely associated with age, BMI and smoking initiation and positively associated with statin use in both samples, with some evidence of larger effect sizes in the CAD sample for age (Int P=0.0064), statin use (Int P<0.001) and BMI (Int P=0.011). In contrast, we found some evidence that the CAD PRS is associated with increased SBP in those without CAD, but this association was largely attenuated in the prevalent CAD sample (Int P=0.020). Similarly, the direction of effect estimates differed between the two samples for type II diabetes with some weak evidence of heterogeneity (heterogeneity P=0.053) (**Table 2**) (**Supplementary Figures 1-7**).

**Table 2.**
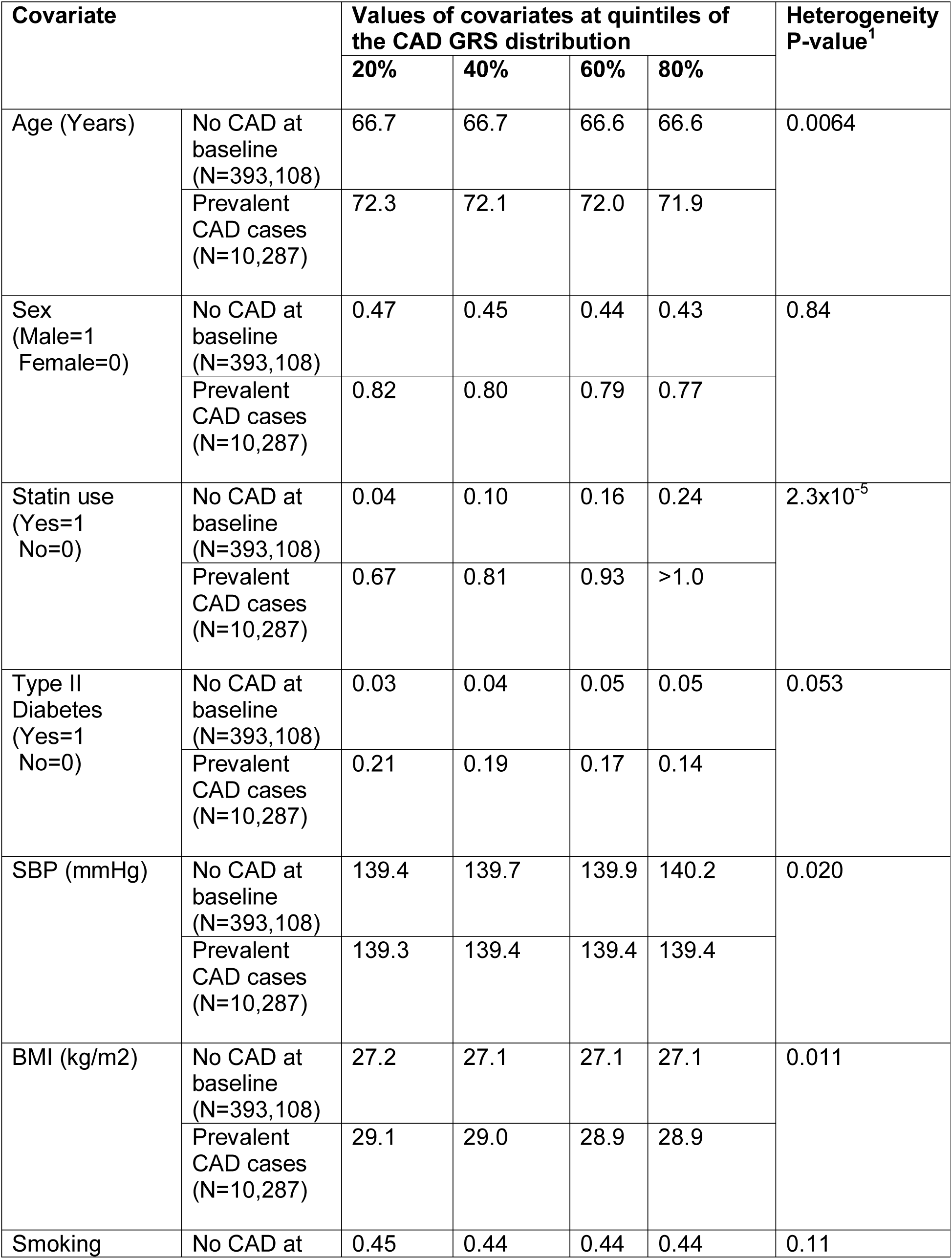
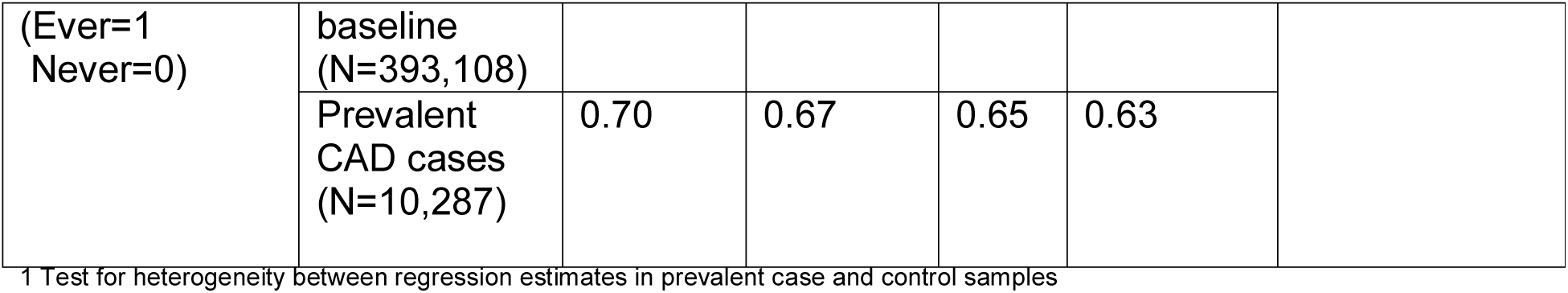
Associations between CAD GRS and covariates.

### Accounting for index event bias

First, we included CAD risk factors (SBP, BMI, smoking, diabetes) and statin use as covariates in the model to account for potential index event bias. Although estimates in general moved slightly closer to the non-case estimates from **Table 1**, we did not find evidence of discernible statistical differences when including these covariates (**Table 3**).

**Table 3:**
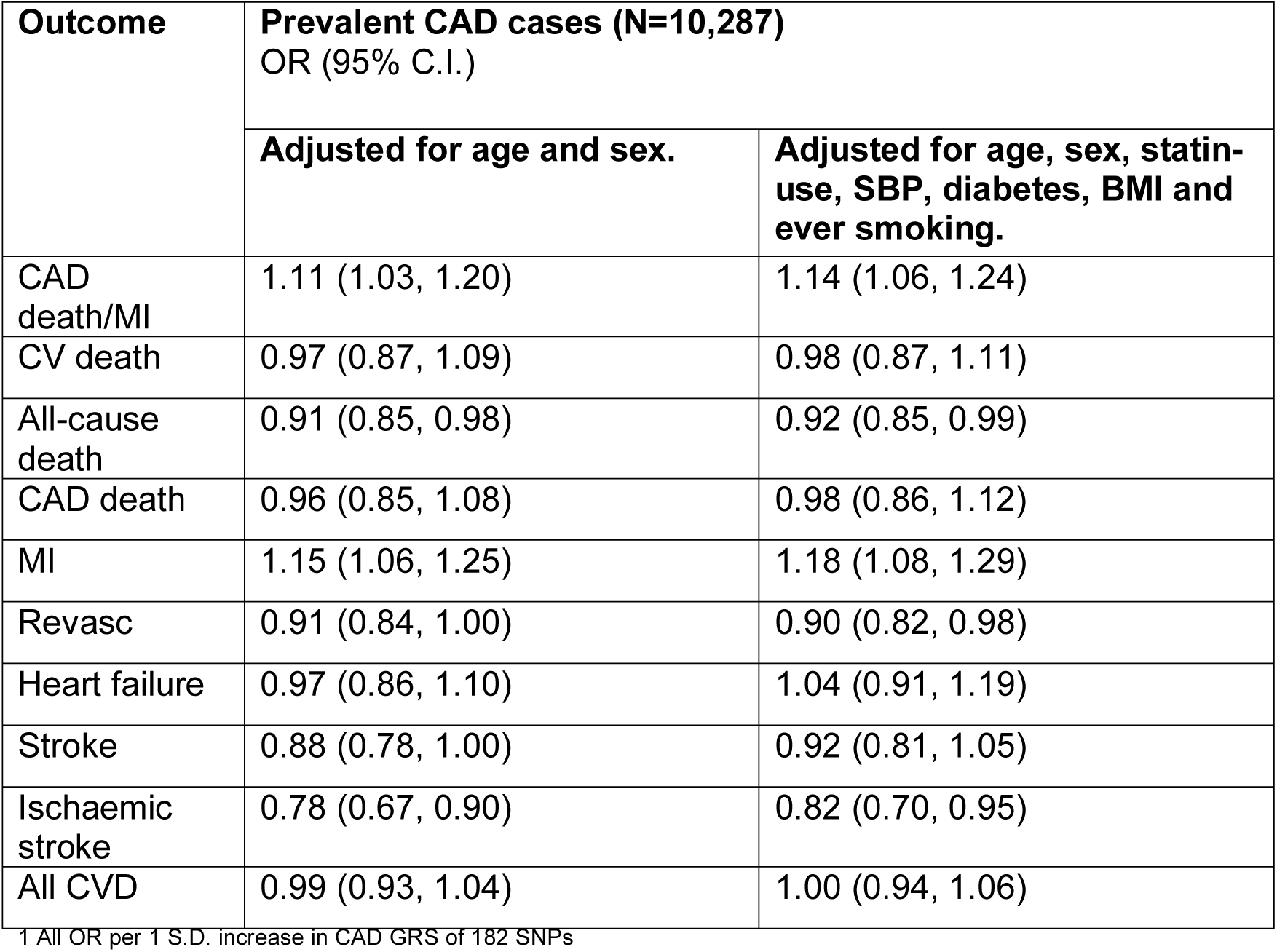
Association of CAD GRS with subsequent incident events adjusted for covariates.

| Outcome | Prevalent CAD cases (N=10,287)<br>OR (95% C.I.) |  |
| --- | --- | --- |
|  | Adjusted for age and sex. | Adjusted for age, sex, statin-use, SBP, diabetes, BMI and ever smoking. |
| CAD death/MI | 1.11 (1.03, 1.20) | 1.14 (1.06, 1.24) |
| CV death | 0.97 (0.87, 1.09) | 0.98 (0.87, 1.11) |
| All-cause death | 0.91 (0.85, 0.98) | 0.92 (0.85, 0.99) |
| CAD death | 0.96 (0.85, 1.08) | 0.98 (0.86, 1.12) |
| MI | 1.15 (1.06, 1.25) | 1.18 (1.08, 1.29) |
| Revasc | 0.91 (0.84, 1.00) | 0.90 (0.82, 0.98) |
| Heart failure | 0.97 (0.86, 1.10) | 1.04 (0.91, 1.19) |
| Stroke | 0.88 (0.78, 1.00) | 0.92 (0.81, 1.05) |
| Ischaemic stroke | 0.78 (0.67, 0.90) | 0.82 (0.70, 0.95) |
| All CVD | 0.99 (0.93, 1.04) | 1.00 (0.94, 1.06) |

<sup>1</sup> All OR per 1 S.D. increase in CAD GRS of 182 SNPs

Second, we applied a method to correct for index event bias in GWAS (32). The regression of genetic effects for prognosis on those for incidence generated a positive slope estimate using both SIMEX (0.0655; 95% C.I. 0.646, 0.0664) and the Hedges-Olkin estimator (0.0516). Prior to the adjustment using the SIMEX estimate, a 1-unit odds increase in genetic liability to CAD was associated with reduced odds of mortality (OR 0.76; 95% C.I. 0.65, 0.90; P=0.0018), directionally concordant with the individual level data analysis which was scaled differently (see **Table 1**). After correction, the association between the CAD PRS and mortality increased in magnitude with a slightly more extreme inverse association, although confidence intervals overlap before and after correction (OR 0.72; 95% C.I. 0.60, 0.85; P=0.0001).

## Discussion

In this study, we have demonstrated that associations of CAD PRS with covariates and incident cardiovascular and fatal outcomes differ between those with and without prior CAD. Notably, we found that associations of the PRS with risk of future MI and CAD death were greatly attenuated among those with established CAD, with some evidence of a positive association for MI but very weak evidence for a positive association with CAD death, compared to those without CAD. Furthermore, we found evidence for inverse associations between the CAD PRS and all-cause death and ischaemic stroke amongst cases which were not present in individuals without known CAD.

These findings could be partially explained by index event bias, whereby stratifying on case status induces non-causal associations between genetic variants and risk factors. For example, individuals with high genetic risk for CAD may develop coronary disease despite low levels of conventional CAD risk factors such as smoking and adiposity. Indeed, we found some evidence that higher CAD PRS are associated with reduced BMI and smoking initiation in both samples, with more extreme effect sizes observed in the case sample suggesting that attenuated associations may be attributable to cases with higher genetic risk being otherwise healthier. Similarly, another possibility is that bias may be induced by the selection of study participants into UK Biobank; individuals with high genetic risk for CAD may be more likely to die prior to being recruited into the study or decline participation for a health reason. This possibility is supported by the inverse association between the CAD PRS and age, again with a larger effect size amongst cases which suggests that individuals with higher genetic risk for CAD may have increased mortality. A further possibility is that the difference in associations are partially explained by aetiological heterogeneity between CAD onset and progression, characterised by differential drivers of stable and unstable plaque risk. However, it seems unlikely that the observed protective associations of the CAD PRS with all-cause death and ischemic stroke are explained by biological differences.

Medication use such as statins may also have contributed to the inverse associations in individuals with prevalent CAD, with previous evidence suggesting that statin use is more effective in those with higher genetic risk to CAD (19, 37). This interaction likely relates to genetic overlap between CAD and LDL cholesterol, the target of statins, with higher genetic risk individuals more likely to have elevated LDL cholesterol. However, although statins may be more effective in individuals with higher genetic risk that doesn’t necessarily equate to lower absolute risk amongst individuals with elevated genetic risk, as our results imply for several outcomes. Indeed, in one of the previously cited studies (19), individuals with higher genetic risk were found to have increased mortality.

To investigate the potential effects of index event bias on our analyses, we applied two distinct methods. However, the two methods shifted estimates in opposite directions; adjusting for covariates moved the estimates towards the non-case sample estimates while the index event correction strengthened the inverse association between the CAD PRS and mortality amongst cases. One possible explanation for the increased inverse association after the index event correction, is that the method assumes that the direct effects of prognosis and incidence are independent. In the context of coronary disease, there are clearly factors which influence both incidence and prognosis, such as LDL cholesterol, suggesting this assumption may not hold.

Our findings have several important implications. First, although we did not formally evaluate prediction metrics, the modest odds ratios observed suggest that despite PRS positively associating with MI risk amongst diseased cases, existing PRS are likely to have limited effectiveness for prediction of subsequent events and therefore risk stratification in this setting (38). These findings imply that genetic prediction of subsequent coronary disease events is likely to require dedicated GWAS of coronary disease progression. Second, our findings contribute to the existing literature (13, 31, 32, 39, 40) emphasising the caution required when using genetic data to infer causality in the context of disease progression. Genetic associations are generally thought to be causal by analogy with Mendelian randomization (41) because of the reduced possibility of confounding and reverse causation, but the observed protective associations of CAD PRS with mortality and ischemic stroke suggest that this may not hold for case-only studies. Index event bias has been shown to have modest impact on individual SNPs effects (39) but our results illustrate that bias likely accumulates when combining multiple markers together in a PRS, which could also affect Mendelian randomization studies. Third, the observed associations between the CAD PRS and age across both CAD cases and controls in UK Biobank suggest that potential bias from selection into UK Biobank requires further investigation and consideration in genetic studies (10).

Our study has notable limitations. First, our analyses used only the UK Biobank and require independent replication in different datasets and populations. Second, we could not differentiate between the effects of possible biases and genuine biological differences between onset and progression. Third, available biomarker data including LDL cholesterol was not available in UK Biobank at the time of writing, so we were unable to explore associations between the CAD PRS and CAD related biomarkers. Fourth, other researchers have derived more accurate PRS from the CardioGramPLUSC4D data than ours (5, 6); however, individual risk prediction was not our goal, and given the positive association of our PRS with CAD incidence we expect the same qualitative findings would result from PRS including a greater number of weakly associated SNPs.

In conclusion, we have illustrated that associations between CAD genetic risk variants and cardiovascular outcomes differ when examined in those with and without prior CAD. This may be due to index event bias, although other possibilities need to be explored. Future work, such as dedicated GWAS of disease progression, by initiatives such as the GENIUS-CHD consortium (40) will aim to further explore genetic differences between onset and progression of CAD.

## Data Availability

The summary statistics for the GWAS of all-cause mortality amongst CAD cases in UK Biobank conducted in this study will be made publicly available upon study publication.

## Acknowledgements

This work was supported by a British Heart Foundation Intermediate Fellowship (Dr Patel, grant number FS/14/76/30933). This research was also supported by the National Institute for Health Research University College London Hospitals Biomedical Research Centre; Dr Schmidt is funded by a British Heart Foundation grant number PG/18/5033837. Prof Hingorani is a National Institute for Health Research Senior Investigator; Prof Asselbergs is supported by University College London Hospitals National Institute for Health Research Biomedical Research Centre, European Union/European Federation of Pharmaceutical Industries and Associations Innovative Medicines Initiative 2 Joint Undertaking BigData@Heart grant n° 116074, the European Union’s Horizon 2020 research and innovation programme under the ERA-NET Co-fund action N°01KL1802 (Druggable-MI-gene) jointly funded by the Dutch Heart Foundation and Netherlands Organization for Health Research and Development (ZonMw). The funder(s) of the study had no role in study design, data collection, data analysis, data interpretation, or writing of the report.

## Data availability

The summary statistics for the GWAS of all-cause mortality amongst CAD cases in UK Biobank conducted in this study will made publicly available upon study publication.

